# Trans-sectoral patient pathways in urgent and emergency care: a Study Protocol for a prospective mixed-methods study in Germany (TRANSPARENT Study)

**DOI:** 10.1101/2025.11.26.25341063

**Authors:** Jonas Bienzeisler, Miriam K. Hertwig, Hauke Heidemeyer, Mohamed Alhaskir, Raphael W. Majeed, Alexander Kombeiz, Wiliam Hoy, Simon Hüning, Franziska Göttgens, Jenny Unterkofler, Susanne Rademacher, Despina Panagiotidis, Viktoria Marewski, Anja Sommer, Wiebke Schirrmeister, Felix Walcher, Ronny Otto, Saskia Ehrentreich, Harry H. Beyel, Viki Peeva, Christopher T. Schwanen, Marco Pegoraro, Beate Zoch-Lesniak, Johannes Pollmanns, Ralf Wittmar, Dominik von Stillfried, Rainer Röhrig, Stefan K. Beckers, Wil M. P. van der Aalst, Jörg C. Brokmann

## Abstract

**Introduction:** Urgent and emergency care in Germany is delivered across multiple, loosely connected sectors. In the absence of coherent, time-resolved data on patient movements between Emergency Medical Services (EMS), out-of-hours primary care, emergency departments (EDs), and inpatient care, inefficiencies and coordination gaps remain difficult to quantify. A process-centric, trans-sectoral analysis is required to characterise real-world patient pathways and identify actionable levers for improvement. The study aims to reconstruct, model, and analyse patient pathways for urgent health complaints across all relevant sectors of the healthcare system in a German model region.

**Methods and analysis:** We will employ a mixed-methods observational study design. Routine data from EMS, out-of-hours primary care, EDs, and subsequent inpatient care will be pseudonymized at source, linked via a trusted third party, and analysed within a trusted research environment. Time-stamped event logs will support process mining for discovery, conformance, and performance analysis alongside descriptive statistics with stratification by context, such as setting, time of day, urgency, and patient cohorts. Anonymous cross-sectional surveys of patients and frontline professionals, complemented by quarterly snapshot surveys in out-of-hours primary care and interviews, will provide convergent evidence on the motives, barriers, and coordination of utilisation behavior. Enrolment for surveys is anticipated from the 4^th^ quarter of 2025; routine data capture covers 1 January–31 December 2026; analyses and dissemination run until 31 December 2027.

**Strengths and limitations:** The multiprofessional, trans-sectoral mixed-methods design with triangulation of perspectives, together with comprehensive routine data spanning the acute-care continuum, provides a robust basis to reconstruct time-resolved pathways and validate findings. Limitations include reliance on routinely collected electronic health records and administrative or billing data, which have variable completeness and coding quality, potential misclassification, and structurally induced missingness. Additionally, constraints arise from record linkage across sources, and the exploratory, observational nature of the analyses limits causal inference.

## Introduction

Fragmentation of urgent and emergency services across disconnected providers results in uncoordinated patient care and inefficient use of resources^1^ ^2^. This protocol presents a mixed-methods study that will integrate linked routine care data with concurrent patient and stakeholder surveys to quantify and characterise time-ordered patient pathways across the urgent and emergency care sectors of a model region in Germany. By reconstructing these real-world sequences and combining them with the perspectives of patients and healthcare professionals, the study will evaluate process efficiency, identify barriers to coordinated care, and generate evidence to support informed navigation of patients within the system.

### Background and Rationale

Urgent and emergency care is a core function of all healthcare systems^2–4^. In many countries, such as Germany, the United States, and the Netherlands, patients themselves decide what constitutes an emergency. They are free to seek help through emergency medical services (EMS), emergency departments (EDs), or primary care providers, including out-of-hours services. Access frequently occurs without prior triage or centralised guidance^5^ ^6^. Episodes with urgent health problems frequently span multiple sectors, yet routine data rarely capture these cross-sectoral movements in a structured or interoperable way ^7^. As a result, it is often unclear what care has already been provided, which referral pathways were involved, and what happens after discharge^8^. The lack of continuity in documentation hinders coordinated care delivery, compromises patient safety, restricts quality monitoring, and undermines strategic planning in healthcare^9^.

The German urgent and emergency care system reflects these challenges in a particularly pronounced way. It is structurally fragmented and regionally variable in both governance and access. Care is delivered across three formally distinct sectors: outpatient care, including out-of-hours service coordinated by the Associations of Statutory Health Insurance Physicians, EMS organised by municipalities, and inpatient care, including emergency departments operated by publicly or privately funded hospitals. Each sector operates under separate responsibilities, funding mechanisms, and data infrastructures^10^. There is no strict gatekeeping system, and patients can access all sectors directly and independently, often without prior triage or coordination^11^. Primary care by general practitioners and specialists during office hours absorbs a large share of demand, while regionally organised out-of-hours primary care provides urgent care at other times. In some regions, primary care is embedded within or closely connected to hospital-based emergency care through co-located out-of-office practices, shared triage protocols, and formal redirection pathways. Access to urgent and emergency care may occur by self-presentation, provider referral, or via two hotlines: 116117 for non-life-threatening problems, and 112, the EMS hotline with regional control centers, for life-threatening emergencies (Figure 1).

**Figure 1:**
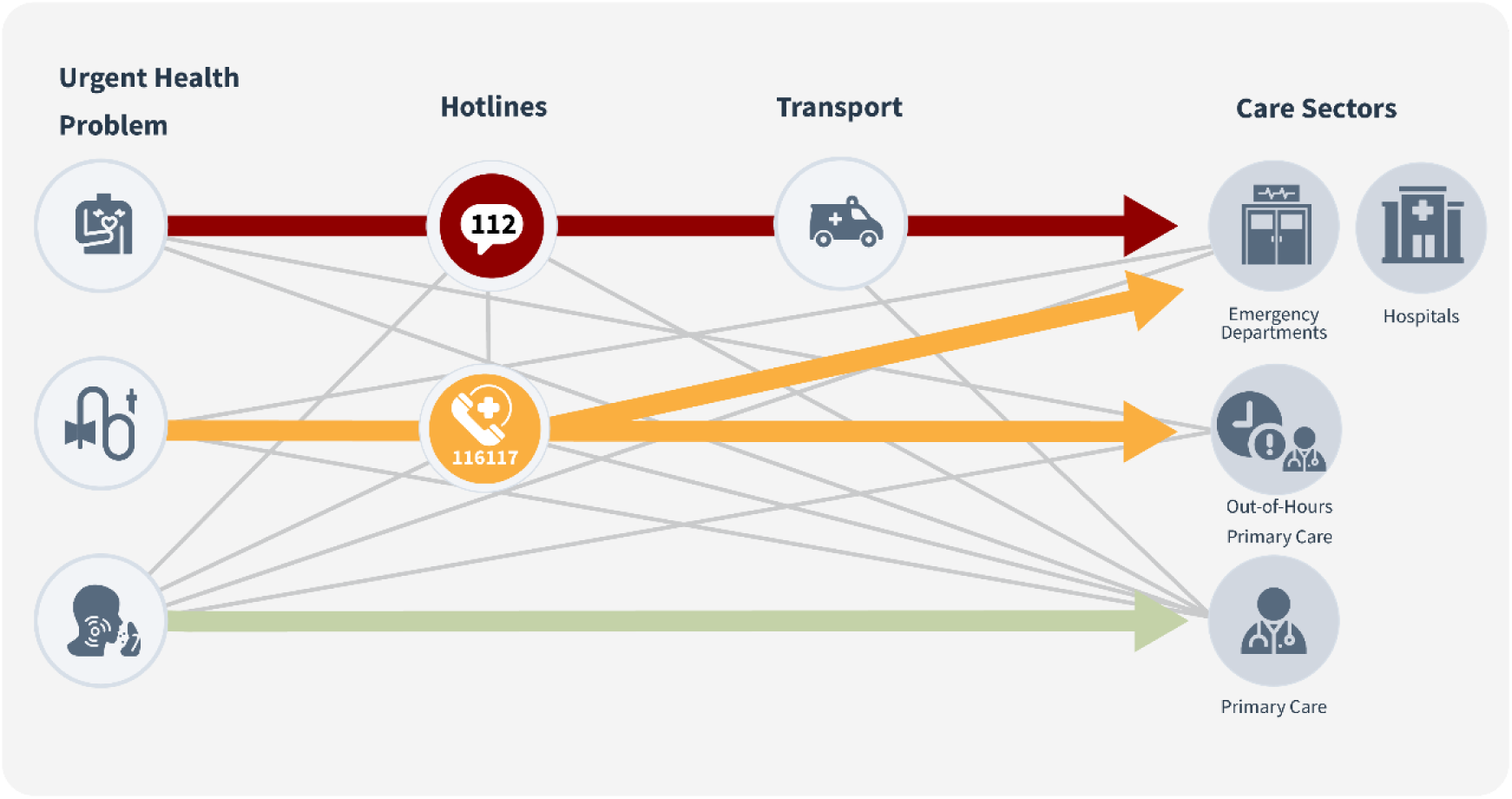
Access pathways for urgent and emergency care in Germany (right). The diagram summarizes typical patient pathways: the hotline 112 (red) for emergencies—dispatch via the municipal control center and EMS transport to an ED (co-located with a hospital); hotline 116117 (yellow) for non-life-threatening out-of-hours problems—telephone triage with referral to out-of-hours services or, as needed, to an ED; and self-presentation/provider referral (green/grey) to primary care or ED. Non-urgent problems are generally managed in primary care (green). 116117 does not dispatch ambulances. Paths are simplified and non-exhaustive; patients may access services directly at multiple entry points.

Reliable patient-level data on intersectoral pathways and process indicators remain scarce in Germany. Electronic health records (EHRs) from urgent and emergency care are often non-standardised, incomplete, non-linkable, especially due to the non-existence of an overarching Unique Patient Identifier in Germany, or inaccessible, limiting robust monitoring, national oversight, and international comparability^12^ ^13^. Importantly, this is not an exclusively German problem. Even in countries with more integrated models, such as the United Kingdom, where the National Health Service has heavily invested in nationwide data infrastructures, real-time visibility into patient trajectories across primary and emergency care remains limited in practice ^14–16^. In Germany, at least partial progress against systemic fragmentation has been achieved through the nationwide AKTIN infrastructure (Alliance for Information and Communication Technology in Intensive Care and Emergency Medicine), which enables access to interoperable EHRs from EDs^17^.

Patients encounter this same fragmentation, often experiencing urgent care as poorly coordinated^18–20^. These perceptions are especially pronounced among individuals with limited health literacy, who may struggle to navigate care options or interpret guidance. Nearly half of European adults demonstrate problematic or inadequate levels of health literacy^21^, a factor consistently associated with increased healthcare utilisation and costs^22^. Limited health literacy contributes to lower uptake of preventive services, more frequent use of urgent and primary care, and a higher risk of avoidable complications^23^. As a result, individuals with low health literacy are disproportionately likely to present to EDs for conditions that could have been managed in primary care^24–27^. Such patterns are frequently labelled as inappropriate, yet they often reflect structural barriers to timely, coordinated, and qualitatively appropriate care rather than intentional misuse^28^ ^29^. Analyses of German data on EMS and primary care use have recently confirmed these assessments: patients with complex needs, who often use primary care, show higher EMS utilisation, resulting in a lack of care continuity^30^. Demographic change will further compound this problem, as ageing populations with complex needs are especially vulnerable to fragmented pathways^29–31^. Navigation behaviour is furthermore shaped by socio-demographic factors such as migration background, health literacy, and the absence of precise steering mechanisms^32^. While digital tools such as symptom checkers have been introduced to improve orientation, their impact on ED utilisation remains limited^33^.

These structural inefficiencies and socio-demographic challenges place additional strain on systems already operating near capacity. Globally, urgent and emergency care is under sustained pressure, with demand rising faster than staffing, bed capacity, and infrastructure expansion^34^. In Germany, EMS operations increase by around 4% annually^34–36^. Ambulatory care–sensitive conditions continue to rise despite the availability of primary care services, with back pain and soft tissue disorders being particularly affected^20^ ^37^. Urgent and emergency care must therefore be understood as a network of interdependent providers whose performance is determined by the sequence and timing of each contact. Whether a patient calls EMS or is first assessed in a triage area influences both resources consumed and outcomes achieved^38^. These pathways remain poorly mapped^39^.

Process-based analyses underscore this potential. Even partial reconstructions of patient pathways have revealed inefficiencies such as diagnostic delays, discharge bottlenecks, and misaligned staffing that become visible only once end-to-end data are available^1^. Process mining is a data-driven method that reconstructs processes from time-stamped records of care activities, enabling the presentation and analysis of pathway structures, sector transitions, and timing in urgent and emergency care^39^ ^40^.

These findings highlight a broader insight: poor system performance is not inevitable, but rather the consequence of care processes that remain opaque^40^ ^41^. When data stops at institutional boundaries, handoffs and resource decisions rely on incomplete information. With coherent, time-resolved data, actionable opportunities for improvement become visible. In Germany, this potential remains largely untapped due to the absence of a unified infrastructure linking EMS, outpatient, and inpatient care. Despite high expenditure and broad coverage, the system lacks mechanisms to map and evaluate cross-sector patient pathways systematically. This study addresses that gap. By combining linked routine data with patient and stakeholder perspectives, it seeks to establish a structural analysis of real-world urgent care pathways. The aim is thus to identify process-related inefficiencies, inform targeted interventions, and strengthen navigation across the urgent and emergency care continuum.

### Objectives

The study aims to generate a comprehensive, process-oriented understanding of patient pathways in urgent and emergency care within a model region in Germany. A patient pathway in this study is defined as the time-stamped sequence of all care events, encounters, and individual traverses during a single episode of an urgent health problem, from initial contact to final treatment. The primary objective is to analyse how adult patients with urgent complaints move across healthcare system sectors in terms of sequence, timing, sector transitions, and frequency.

Secondary objectives are to assess how well comprehensively linked routine data can reconstruct patient pathways and how these data-driven reconstructions align with patient- and provider-reported experiences, decisions, and perceived care processes. The study will further identify subjective and structural factors such as knowledge gaps, accessibility issues, or navigation uncertainty that are associated with potentially inefficient care use and will derive strategies for patient-centred guidance and system-level steering. In addition, pathway variants associated with multiple, redundant, or potentially inappropriate utilisation will be examined to build a valid basis for developing predictive indicators for repeated use or avoidable escalation across care sectors. The study will also explore the role of pain as a frequent and clinically relevant presenting complaint in urgent care and derive targeted improvement strategies within this subgroup, particularly with respect to timely and appropriate navigation.

The resulting insights will serve as the empirical foundation for patient-centred guidance, system optimisation, and the development of targeted empowerment strategies. Furthermore, the study will translate its findings into plain-language materials and data-driven recommendations to strengthen both patient care and health literacy of the model regions’ population. The study will contribute to the establishment of a sustainable infrastructure for research, quality assurance, and public health surveillance by extending the nationwide AKTIN infrastructure to include prehospital data streams, providing robust indicators to support quality reporting and future health services research.

## Methods and Analysis

This study will integrate quantitative real-world data from urgent and emergency care with survey and interview evidence. Patient and public involvement will ensure that priorities, interpretation, and dissemination are aligned with patient needs and care realities. Interests will be represented in a project advisory board including patient organisations and professional associations, such as the German Coalition for Patient Safety and the German Interdisciplinary Association for Intensive Care and Emergency Medicine.

### Study Design

The study will follow a mixed-methods observational design (Figure 2). The quantitative core is based on pseudonymized, time-stamped records from all relevant sectors in a model region, linked at the case level through a trusted third party. Event logs derived from these data will be used to reconstruct complete care episodes and to identify pathway variants, throughout time, and indicators of inefficiency. To complement and contextualise these analyses, anonymous cross-sectional surveys of patients and healthcare providers as relevant stakeholders will capture motives, barriers, and decision-making processes during an urgent health problem episode, while quarterly snapshot surveys of out-of-hours services will provide validation. Stakeholder interviews conducted after preliminary model development will further support the interpretation of the developed process models and highlight system-level constraints.

**Figure 2:**
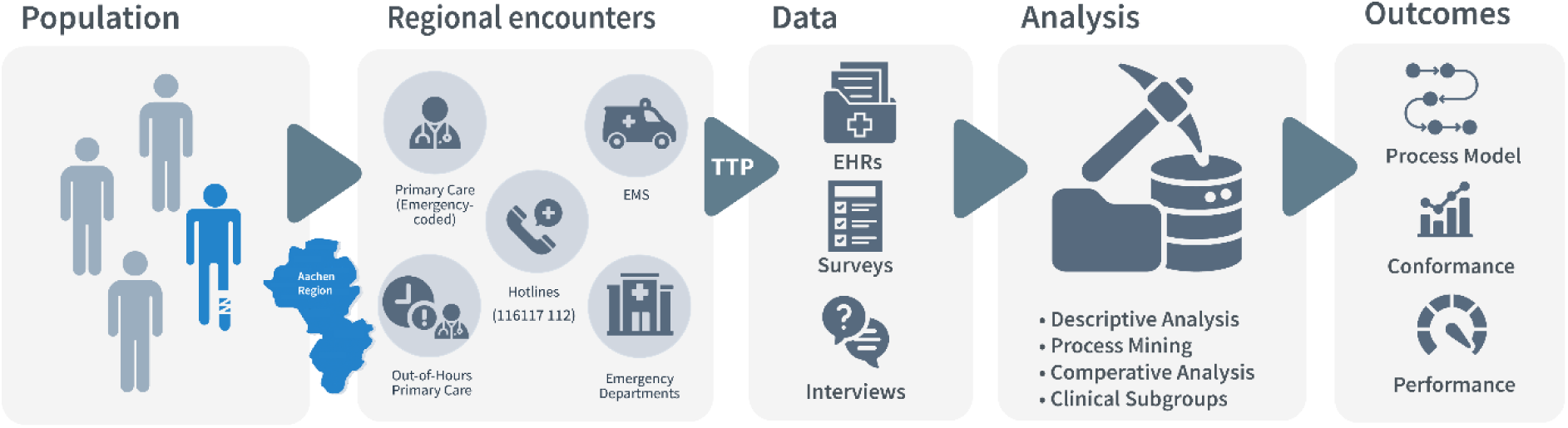
Study design. The study population consists of individuals who had an urgent or emergency encounter in the city of Aachen and its surrounding municipalities during the observation period. Eligible encounters include Emergency Departments, Emergency Medical Services (112 hotline), out-of-hours primary care (116117 hotline), and primary care visits coded as an emergency. Routine EHRs from these sectors are pseudonymized at source and linked by a trusted third party (TTP). The harmonised dataset is transformed into time-stamped event logs for process mining, generating process maps, conformance and performance metrics, and pathway indicators. Surveys (patients and providers) and stakeholder interviews run in parallel to contextualise findings and support validation.

### Study Setting

The study will be conducted in the region of Aachen in western Germany with approximately 582,000 inhabitants. It comprises the independent city of Aachen with around 263,000 residents and the surrounding municipalities. The region is located near the borders with Belgium and the Netherlands and combines urban centres with semi-rural communities (Figure 3). It is served by seven hospitals with emergency departments, including University Hospital RWTH Aachen (tertiary care), Rhein-Maas Klinikum (specialist and urgent care), and an additional five general hospitals; together, these institutions account for more than 150,000 ED visits per year.

**Figure 3:**
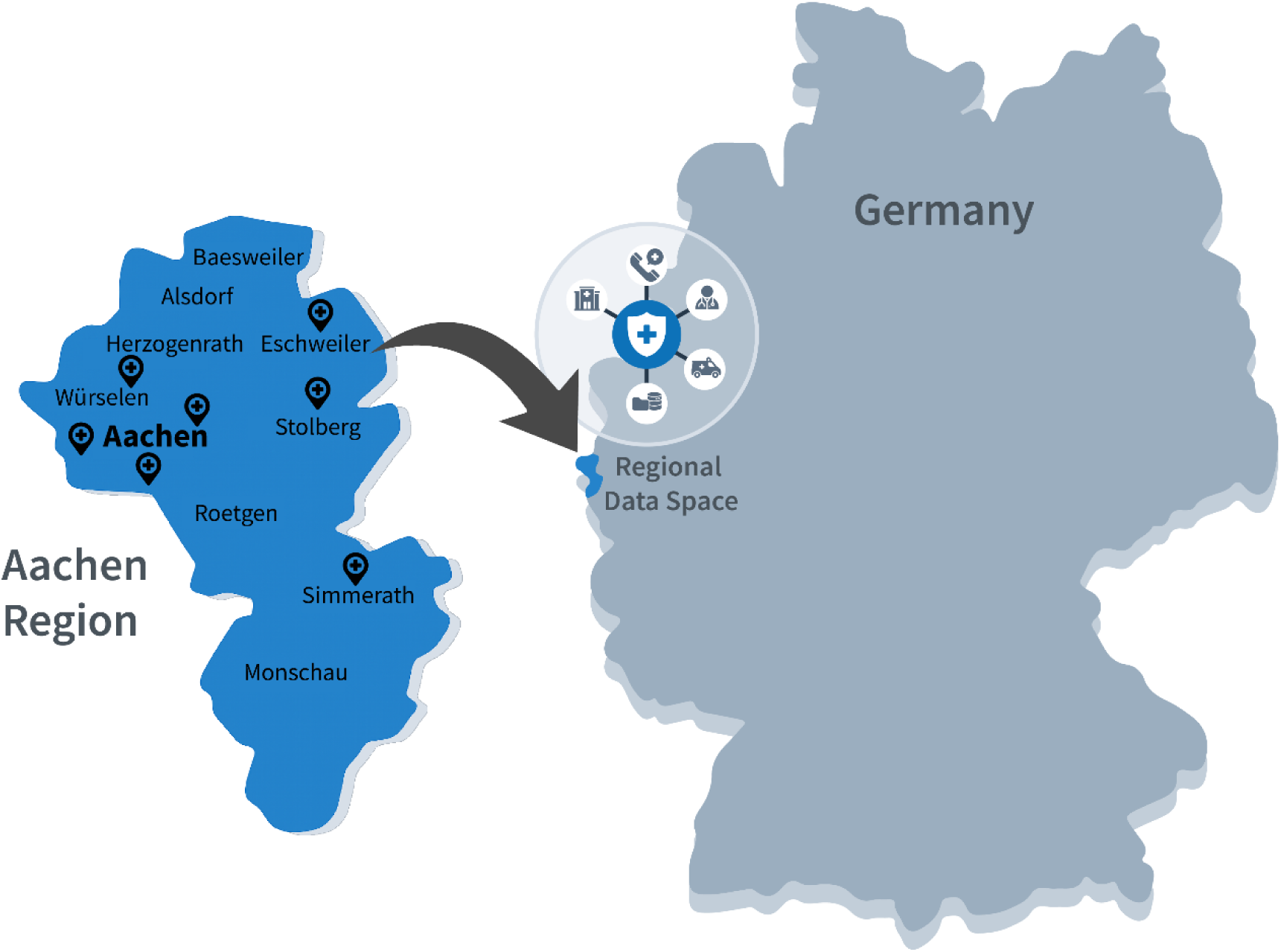
Schematic overview of the Aachen region and its location in Germany (not to scale). The highlighted area represents the model region in which a dedicated data space for urgent and emergency care is being established, enabling event-log analysis of patient pathways across sectors.

Outpatient and out-of-hours services are coordinated by the Association of Statutory Health Insurance Physicians North Rhine, which registers all office-based general practitioners and specialists, organises ambulatory care during office hours and structured out-of-hours coverage, and operates the nationwide hotline 116117 for non-life-threatening problems with telephone triage and referral. As of August 2025, the study region includes eight out-of-hours practices (four general, one ophthalmology, two paediatric, one ear-nose-throat) coordinated by the Association of Statutory Health Insurance Physicians North Rhine, each staffed in rotation by around 25 to 215 physicians, along with roughly 377 full time equivalents as general practitioners and around 720 full time equivalents as specialists operating in primary care. They provide service during evenings, nights, and weekends. Two of the out-of-hour practices are co-located with general hospitals.

Most EMS missions, approximately 110,000 cases per year, are coordinated by the integrated control centre in Aachen, responsible for fire protection, disaster coordination, and helicopter dispatch. A smaller control centre in Eschweiler manages part of the fire and rescue missions in its immediate vicinity. EMS is delivered jointly by professional fire departments and non-profit aid organisations, such as the German Red Cross, operating multiple ambulance stations and physician-staffed emergency vehicles to ensure a 24/7 response capacity.

### Eligibility Criteria

The quantitative core comprises all patients who had an urgent or emergency care encounter in the study region during the observation period. An urgent or emergency encounter is defined, based on established clinical and health services research definitions, as a situation involving sudden or progressive physical or psychological deterioration that prompts the affected person or a third party to seek immediate medical attention^42^. This includes both time-critical, life-threatening emergencies and urgent presentations of acute or chronic conditions with risk of deterioration or complications. For this study, an encounter is eligible if it is (i) a documented contact with any urgent or emergency care service (medical hotline 116117, dispatch centres approached via 112 emergency hotline, EMS, out-of-hours primary care, or hospital EDs) or (ii) a primary-care consultation with a general practitioner or specialist which is coded as an emergency, within the observation period. Surveys will be conducted anonymously via digital platforms, with recruitment through on-site materials or organisational distribution channels. Eligibility requires an encounter with the urgent and emergency care system in the study region, either as a patient (or as reported by parents or guardians for minors) or as a stakeholder working within the regional care structures. Interview partners will also be recruited from the study context to ensure that their perspectives reflect the local organisation of care.

### Data Sources and Collection Procedures

The study will integrate three parallel data streams: EHR data from various sectors, anonymised patient and healthcare professional surveys, and qualitative interviews with stakeholders.

### Electronic Health Records

The primary stream of routine care data combines prehospital, hospital, and outpatient encounters into unified, time-stamped patient pathways. Clinical data from hospital EDs are obtained through the AKTIN infrastructure, to which all EDs in the region are being connected. The AKTIN infrastructure is a nationwide, federated data network operated within the Network University Medicine^43^. The infrastructure enables the continuous and standardised re-use of EHRs following the DIVI emergency department dataset collected within ED information systems^44^ ^45^. Data includes routine documentation and, where applicable, will be complemented by discharge data (diagnoses, procedures, length of stay). The infrastructure also supplies structural parameters (e.g., speciality coverage, triage system, etc.).

Prehospital data are provided by the regional emergency dispatch centres, which document all incoming 112 calls. Each call is recorded in a standardised dispatch protocol that includes timestamps, dispatch priorities, vehicle deployment, and information about geospatial movement. EMS personnel complete structured handover reports in line with the DIVI-MIND2 standard during service, covering patient history, vital signs, prehospital interventions, and time of transfer to hospital care. These reports are currently handwritten but will be sequentially transformed into digital documentation, which can then be digitally transmitted to the EDs as well as the AKTIN infrastructure and enable precise linkage between prehospital and in-hospital care events^46^.

The Association of Statutory Health Insurance Physicians North Rhine will provide complementary outpatient data. This includes routine claims data under § 295 of the German Social Code, Book V (SGB V), covering urgent cases treated by contracted general practitioners and specialists, as well as ambulatory emergency care provided during both regular hours and organised out-of-hours services. Data from the 116117 medical hotline will also be provided and will include age, gender, timestamps, the call taker’s decision regarding further care, and, in most cases, data of the Structured Medical Initial Assessment in Germany (SmED) protocol, which contains the medical problem and a recommendation regarding the urgency and the appropriate level of care^47^.

### Surveys

Anonymous cross-sectional surveys will be conducted targeting patients and stakeholders from urgent and emergency care. The instruments are designed to capture self-reported experiences, perceived barriers, and decision-making across the urgent care continuum. Questionnaires for patients and ED employees were developed based on existing instruments identified through a systematic literature review, with candidate items appraised using established criteria for clarity and measurement validity. In the absence of validated instruments, questionnaires for EMS and out-of-hours primary care will be developed specifically for this study based on the literature and expert input, cognitively pretested, and evaluated for content validity and reliability before deployment. Questionnaires will be version-controlled, and any refinements following pretesting will be documented in a change log. The final instruments, together with the underlying literature review and documentation of the development process, will be published alongside the study results to ensure transparency and reproducibility.

The surveys for patients and ED personnel will entail closed-ended items, enabling standardised measurement. Survey for out-of-hours primary care providers and EMS personnel will combine closed-ended items with selected open-ended questions to capture context-specific experiences and coordination-related constraints. Patient questionnaires will be distributed via QR codes displayed in participating EDs and out-of-hours practices and can be completed on-site during waiting times. Healthcare professionals will be invited through personalised links distributed via institutional mailing lists and contacts provided by medical directors of the participating providers. To ensure feasibility across settings, the instruments will be designed for completion within approximately ten minutes and optimised for use on mobile devices.

The study will further include quarterly snapshot surveys of primary care in 2026. On pre-specified reference dates in each quarter, general practitioners will be contacted directly by email and asked to complete a short, standardised questionnaire for all contacts seen that day. It will capture aggregate counts and minimal demographics, reasons for encounter, diagnoses (ICD-10-GM), and, where applicable, urgency, services provided, and referrals to EMS or EDs. To support contextual validation, the nationwide primary care claims dataset (under § 295 SGB V) will be analysed separately. These reference data will serve as a benchmark but will not be linked to individual patient records or survey responses.

### Interviews

Stakeholder interviews will be conducted after preliminary process models have been discovered, to validate findings and contextualise system-level constraints. The interviews will follow a semi-structured, qualitative design, focusing on the plausibility, completeness, and interpretability of the reconstructed pathways, as well as identifying structural or organisational factors that may not be visible in routine data. Given that the process models and related hypotheses are data-driven, the exact interview guideline cannot be pre-specified; it will be finalised immediately before the interview phase and made publicly available alongside the study results. Interviews will be audio-recorded, transcribed, and analysed thematically, with two independent researchers coding the material to enhance reliability.

### Outcomes

The primary outcome of this study is the reconstruction and quantitative description of trans-sectoral patient pathways for adult patients (aged 18 years and above) with acute complaints in the model region. We operationalise a patient pathway as the time-ordered sequence of urgent or emergency care encounters that occur within a defined temporal window of 72 hours^48^ (with longer intervals examined exploratively) or that share the same main complaint cluster (CEDIS category or leading ICD-10 group). A new episode is assumed if the interval between encounters exceeds this window and the complaint cluster changes, with pathway length limited to 30 days in accordance with existing quality indicators for urgent and emergency care^49^. Longer sequences are considered chronic utilisation patterns. For emergency patients, we define their pathways as the contiguous core segment of such sequences, where successive encounters follow without interruptions longer than 360 minutes and directly address the urgent complaint. The outcome itself is defined as the prevalence and distribution of distinct pathway variants, characterised by their sequence of care encounters, timing, sectoral transitions, and overall patient pathways.

Secondary outcomes extend this analysis to additional dimensions of healthcare utilisation and pathway quality. These include the relative frequencies of patient pathway variants as the empirical spectrum of patient flows; the alignment between reconstructed and reported pathways as an indicator of validity and completeness; and the identification of inefficient or avoidable utilisation, such as repeated emergency contacts, use of high-acuity services for low-urgency complaints like ambulatory care-sensitive conditions or bypassing of coordination structures like the 116117 hotline. A further outcome is the characterisation of non-traumatic pain-related patient pathways, assessing patient patterns and transitions in this clinically relevant subgroup. Reported barriers to access and coordination, including decision-making drivers and navigation deficits, will be captured through structured surveys and analysed in relation to pathway characteristics. As a secondary methodological outcome, the accuracy of record linkage and the performance of the data management infrastructure will be evaluated.

While all encounters, including those of patients under 18 years, are eligible for inclusion, the primary outcome analyses are restricted to adults. Pediatric pathways, especially with recurring encounters, are highly heterogeneous and are suspected to have underlying reasons like rare diseases^50^. Encounters of patients under 18 years will thus be reported separately in exploratory analyses.

### Study Timeline

Routine care data will be collected for the full calendar year 2026, encompassing EMS, EDs, office-based physicians, out-of-hours practices, dispatch centres, and hotline service. Data extraction will follow a harmonised schedule across all participating institutions. The cross-sectional surveys of patients and healthcare professionals will be conducted over 12 months, commencing in the fourth quarter of 2025, following pretesting of the questionnaires and registration of the study protocol. Patients will be invited to participate while present in emergency departments or general practices, using publicly displayed QR codes, typically during waiting periods after registration. Healthcare professionals will be contacted via institutional mailing lists, and quarterly snapshot surveys will be conducted on one predefined day per quarter throughout 2026. Data deliveries are planned on a quarterly basis for quality and plausibility checks; the final analysis will be conducted after database lock on 31 December 2026.

### Sample Size and Power Considerations

Based on historical data from 2022, which serves as the planning basis, we currently estimate the total population to consist of approximately 207,000 ED encounters, 62,488 calls to the 116117 medical hotline, of which 18,746 were classified as acute cases, 70,000 EMS deployments, and 57,388 documented contacts with the out-of-hours primary care service. These historical figures reflect real-world utilisation patterns in the model region but must be interpreted with caution due to potential deviations caused by COVID-19-related healthcare dynamics during that period. These projected sample sizes are sufficient for stratified descriptive analyses and triangulation with routine data; however, they do not follow a formal power calculation, as the study’s mixed-methods design is primarily exploratory in nature.

For the accompanying surveys, we assume a response rate of 10% for patients and 30% for healthcare professionals, based on previous digital, self-administered survey studies. From the available base population, this yields an expected survey sample of approximately 26,400 patients, including around 20,700 from emergency departments and 5,740 from the out-of-hours primary care service. Among the estimated 1,600 healthcare professionals in urgent and emergency care in the region, including 270 staff in EDs, 800 EMS personnel, and 500 physicians in primary care, we expect a total of 470 completed stakeholder surveys (80 from ED staff, 240 from EMS, and 150 from primary care).

### Data Management and Confidentiality

The study utilises pseudonymized routine data and anonymous survey data. Routine data will be sourced from its origin and pseudonymized locally. Direct identifiers are replaced by salted cryptographic hashes. Fields that require approximate matching are encoded with Bloom filter encoding. Pseudonyms and predefined quasi-identifiers are transferred to a trusted third party for privacy-preserving record linkage. The linked and de-identified research dataset is analysed exclusively within the AKTIN Trusted Data Analytics Centre, a protected research environment used within the AKTIN infrastructure. Data processing follows the provisions of the German Health Care Utilisation Act, an opening clause for the General Data Protection Regulation (GDPR), enabling the use of health data for research in the public interest. Hereby, a regional data space for urgent and emergency care is established^51^. Survey data will be collected anonymously via structured digital questionnaires from patients and providers, using separate forms within an online data capture system.

### Statistical and Analytical Methods

For each patient, sequences of contacts will be reconstructed to form event logs representing trans-sectoral patient pathways. The detailed specification of statistical methods and modelling approaches are provided in a separate Statistical Analysis Plan^52^. All statistical tests will be two-sided, with a significance level set at p < 0.05. Effect sizes and 95% CIs will be reported alongside p-values.

### Quantitative Descriptive Statistics

Descriptive statistics will characterise the study population, encounters, and aggregated care episodes, following conventional standards for exploratory health services research. Stratifications will be applied by care sector, demographic subgroups, presenting complaint, urgency level, and care context (e.g. urban vs. non-urban using thresholds from the German Federal Statistical Office or comparable classifications). Demographic and clinical variables will include age, sex, presenting complaints, triage categories, and urgency level. Urban versus non-urban residence will be defined based on postal code and population density. Encounters will be described by type, volume, and temporal distribution. Continuous variables will be summarised using means and standard deviations or medians and interquartile ranges; categorical variables as counts and percentages. Distributions will be presented graphically (e.g. histograms, density plots). Pathway transitions between sectors will be visualised using diagrams such as Sankey plots. Between-site variation and clustering will be addressed using multilevel approaches where appropriate, with fixed or random effects depending on the number of units.

### Process-Modeling

The collected data will be analysed with a focus on care pathways and their underlying process structures using process mining methodologies. Depending on scope and feasibility, various event logs will be created, including sector-specific logs as well as combined datasets to capture duplicates, multiple encounters, or inappropriate utilisation. In addition, data-driven definitions of duplicates, multiple encounters, and inappropriate utilisation will be derived to ensure consistent characterisation across sectors.

Descriptive process characteristics (pathway length, number and type of sector transitions, inter-event intervals, activity distributions, temporal patterns, and process variants) will be reported^53^. Qualitative expert interviews will elicit de jure process models, which will be compared against reconstructed event logs to assess plausibility, completeness, and discrepancies. We will reconstruct pathway models from the event logs using established process-discovery methods. We will evaluate candidate models using conformance metrics. The best-scoring models will be rendered as human-readable diagrams and reviewed with sector experts to confirm plausibility and completeness; the model will then be selected for downstream performance analysis (e.g., inter-event intervals, transition times, bottlenecks). Where models contain decision points, we will apply decision mining^53^ to derive interpretable rules linking case attributes to routing choices. Comparative analyses will test differences in pathway characteristics between subgroups (e.g. by age or urban/rural context), with effect sizes reported alongside p-values. Performance analyses will explore potential bottlenecks.

### Analysis of Survey Data

Survey data will be analysed descriptively and stratified by key subgroups (e.g. age, gender, perceived urgency, reason for attendance). Associations between sociodemographic characteristics, health-related behaviour, and ED use will first be examined using bivariate methods, with effect sizes and confidence intervals reported. Where appropriate, multivariable regression models will be applied to adjust for confounding and identify predictors of frequent or non-urgent utilisation. Cluster analyses may be conducted to identify distinct utilisation profiles. Open-ended stakeholder responses will be analysed using qualitative content analysis to capture recurring themes and system-level issues.

### Mixed-Methods Integration

Quantitative routine data and survey results will be integrated using joint displays to align reconstructed pathways with self-reported experiences and expert interviews. This will highlight concordance, complementarity, or divergence across sources, focusing on aspects such as timing, access, and repeat use.

### Subgroup Analyses

Analyses will be stratified by demographic, clinical, and contextual variables including age, sex, urgency, population density, time of presentation, comorbidity indices, and sector of initial contact. A dedicated subgroup analysis will focus on pain-related complaints. The cohort will be identified based on CEDIS chapter classifications, triage diagrams, and ICD-10 codes. Additional subgroups will be defined by presenting complaints, discharge diagnoses, or sector-specific utilisation patterns. Sensitivity analyses will evaluate the impact of missing data and alternative definitions of pathway windows.

### Handling Missing Data

Missing data are inherent to routine documentation across all sectors. In urgent and emergency settings, records are not collected primarily for research, and gaps are often structurally induced (e.g., undocumented handovers, missing timestamps, system constraints). We therefore do not assume missing at random and treat missingness as potentially informative. Missing values will be retained and described as part of the process characterisation. For time-critical steps (e.g., timestamps, triage assessments), missingness will be profiled by sector and subgroup, and sensitivity analyses will assess its impact on process modelling.

### Bias and Confounding Mitigation

To avoid introducing selection bias into the core dataset of routine care records, individual patient consent will not be obtained for the retrospective use of pseudonymized routine data. Given the heterogeneity and administrative origin of several data sources—particularly billing records from general practitioners and the 116117 out-of-hours service—there is a risk of classification bias, incomplete documentation, and systematic distortions of clinical content or urgency levels. To mitigate these risks, a quarterly Snapshot survey will be conducted across outpatient emergency providers to enable independent validation of service categories, presenting complaints, and triage classifications. This structured, provider-reported survey offers a consensus-based reference to calibrate and interpret routine data across contexts. To address potential misclassification and incomplete documentation in ED data, we will, as feasible, exploratively compare ED records with de-identified data from the AKTIN infrastructure. This nationwide benchmarking will provide descriptive context on documentation patterns and potential systematic deviations.

To further adapt for potential biases and information gaps in routine data, the study employs a triangulation strategy that integrates patient and provider surveys. These surveys are specifically designed to capture subjective decision-making, access barriers, and care experiences that may not be visible in routine documentation. While survey participation itself introduces the possibility of sampling bias, mitigation strategies include the use of multilingual questionnaires, low-threshold digital access and on-site recruitment during care episodes to ensure representation across diverse demographic and linguistic groups.

Confounding related to contextual heterogeneity will be addressed by stratified analyses and, where appropriate, multivariable adjustment. Key pathway characteristics such as process duration, transitions, and repeated contacts will be analysed in relation to these variables.

### Monitoring and Data Quality Assurance

Given the retrospective and pseudonymized nature of the routine care data, no on-site monitoring or individual source data verification will be conducted at the clinical sites. Instead, data integrity and structural validation are ensured through the AKTIN infrastructure, which includes standardised export interfaces, automated data formatting checks, and conformance validation at the local data warehouse level. These technical measures ensure consistent semantic and syntactic data quality prior to extraction. For the cross-sectional surveys, completeness and plausibility checks will be implemented at the point of digital data entry using pre-defined logic and branching rules.

The evaluation of record linkage and data management will be conducted using quantitative accuracy measures, including match rates, sensitivity, specificity, and positive predictive value of linkage algorithms against reference identifiers where available. Data quality will be assessed along established dimensions^54^.

### Ethics and Dissemination

#### Ethics

The study received ethical approval from the Ethics Committee of the Medical Faculty at RWTH Aachen University (EK 25-351). The committee raised no ethical or legal concerns, judged the expected benefits of the planned data processing to clearly outweigh potential risks, and approved the study without requiring revisions. The study will be conducted in accordance with the Declaration of Helsinki^55^, relevant national regulations, and the GDPR. Any substantial amendments to the study protocol, including changes to the design, methodology, or survey instruments, will be submitted to the ethics committee for review prior to implementation. Minor procedural modifications that do not affect participant rights or data protection will be documented in the trial documentation and reported in the final study dissemination.

For the survey modules, written or digital consent will not be collected, as participation is anonymous and of minimal risk. Participation is entirely voluntary. Patients and healthcare professionals will receive prior information about the purpose and scope of the study through on-site materials (e.g., flyers or posters). Upon scanning a QR code, participants are directed to an online platform where an introductory page outlines the study’s objectives, the responsible institution, assurances of voluntary participation, the absence of personal data collection, and the anonymity of responses.

Routine care data will be processed without individual informed consent in accordance with the German Health Care Utilisation Act, which permits the use of pseudonymized routine data for public interest research purposes. This legal framework is explicitly designed to enable analyses of healthcare processes without introducing selection bias that may arise from consent-dependent inclusion. As such, the study adheres to the principle of data minimisation, using only pseudonymized records, with all linkage conducted within a trusted third-party infrastructure.

### Risk–benefit assessment

The resulting evidence of this study will support patient-centered navigation, inform system-level planning, and strengthen quality assurance, thereby directly improving care for future patients. The study inevitably involves the processing of special categories of personal data. Due to the absence of individual informed consent, data subjects’ cannot directly control the processing of their records. Acute and emergency care trajectories are highly individual and may reflect chronic conditions, repeated encounters, or complex patterns of service use or even misuse. Such unique constellations can make patients particularly vulnerable and increase the theoretical possibility of re-identification despite rigorous pseudonymization. Obtaining consent in this context is, however, not feasible. Patients frequently present in urgent or life-threatening conditions, decision-making capacity may be impaired, and trajectories often span multiple institutions, making re-contact for consent impractical or impossible. A consent-based approach would systematically exclude precisely those patients with the most complex and resource-intensive pathways. This would not only bias the dataset but also prevent meaningful insight into the systemic inefficiencies and coordination gaps that affect these vulnerable groups most directly. In contrast, the expected benefits for patients and society are substantial. By enabling the reconstruction of trans-sectoral, time-resolved care pathways, the study addresses critical gaps in the transparency of acute and emergency care delivery. This perspective is indispensable for identifying inefficiencies, avoidable duplications, and safety risks that remain hidden in isolated sectoral datasets. These benefits are realised under strict technical and organisational safeguards, implementing a multi-layered protection concept. All data are pseudonymized at the source, direct identifiers are removed, and quasi-identifiers are secured through privacy-preserving encoding. Record linkage is performed exclusively by a trusted third party, and all analyses are conducted within a trusted research environment, operated under professional confidentiality obligations. Under these conditions, the residual risk for individuals is minimal, while the societal and patient benefit is substantial. On this basis, the public interest in improving emergency care outweighs the risks of data processing under the applied safeguards. The study is therefore lawfully grounded in § 6 of the German Healthcare Data Utilisation Act, which permits the use of pseudonymized routine data for research conducted in the public interest.

### Dissemination and Ancillary Care

No ancillary care will be provided in the context of this observational study. Given the retrospective nature of the routine data analyses and the non-interventional design of the survey modules, there is no provision for direct clinical follow-up or additional support beyond participation.

The results of this study will be disseminated through peer-reviewed publications, conference presentations, and summary reports made available to participating institutions and relevant stakeholders in the healthcare system. Emphasis will be placed on communicating findings that inform sectoral coordination, resource utilisation, and patient navigation in urgent and emergency care. In parallel, insights from the data analyses will be used to develop practical guidance for improving patient orientation within the healthcare system. This includes identifying opportunities to strengthen health literacy and understanding of care pathways among patients with urgent complaints. To this end, existing strategies to support informed care-seeking behaviour will be reviewed and contextualised using study results. Depending on the identified needs, tailored information materials may be developed to communicate the structure and appropriate use of urgent care services. The materials will be written in clear, accessible language to reach patient groups with limited health system knowledge or language barriers, and will be disseminated via peer-reviewed publications, conference presentations, and summary reports for participating institutions and stakeholders.

## Supporting information

Statistical Analysis Plan

SPIRIT Checklist

## Data Availability

All data produced in the present study are available upon reasonable request. Such data from the AKTIN Emergency Department Data Registry cannot be shared publicly; access to de-identified datasets may be granted upon request and following approval by the AKTIN Data Access Committee.

## Author Contributions

Authors’ contribution (CREDIT): Conceptualization: JB, MH, JU, RR, JCB Methodology: JB, MH, HH Software: HH, AK, SH, WH Project administration: JB, MH Visualization: JB Writing – original draft: JB, MH Writing – review & editing: MA, RWM, AK, SH, FG, JU, SR, DP, VM, AS, WS, RO, SE, HHB, VP, CS, MP, BZL, JP, RW, DvS, RR, SB, WVDA, JCB Supervision: RR, SB, WVDA

## Use of artificial intelligence

While preparing this work, the author used Grammarly and OpenAI’s ChatGPT −4 and 5 language models to assist with language refinement in the manuscript. After using these tools, the authors reviewed and edited the content as needed, taking full responsibility for the final content of the work.

