## Supplementary material for "Trans-sectoral patient pathways in urgent and emergency care: a Study Protocol for a prospective mixed-methods study in Germany (TRANSPARENT Study)": Statistical Analysis Plan

### Statistical Analysis Plan (SAP)

| | |
|---|---|
| Study | TRANSPARENT – Trans-sectoral patient pathways in urgent and emergency care |
| Study Registration Number | DRKS00035916 |
| Sponsor | University Hospital RWTH Aachen, Pauwelsstraße 3052074 Aachen, GERMANY |
| Intervention | Health policy intervention: reconstruction and analysis of trans-sectoral patient pathways in acute and emergency care |
| Protocol Number | Protocol v1.0 (November 26th, 2025) |
| Phase | Observational health services research study (not applicable to drug/vaccine phases) |
| Statistical Analysis Plan Version | v1.0 |
| Statistical Analysis Plan Date | September 30th, 2025 |
| Author (Prepared by) | Jonas Bienzeisler, Institute of Medical Informatics, University Hospital RWTH Aachen, Aachen, GERMANY |
| Reviewed by | Ronny Otto, Institute for Public Health in Acute Medicine (IPHAM), University of Magdeburg, Magdeburg, GERMANY |
| Approved by | Jörg C. Brokmann, Centre for Clinical Acute- and Emergency Medicine, University Hospital RWTH Aachen, Aachen, GERMANY |

#### Introduction

This SAP complements the study protocol and provides detailed descriptions of the planned analyses for the TRANSPARENT project. It defines study populations, outcomes, statistical methods, and approaches to sensitivity and subgroup analyses. The SAP ensures transparency, reproducibility, and minimises the risk of bias. Analyses not prospectively defined will be explicitly reported as exploratory.

##### Timing of SAP Development

The SAP was developed in parallel with the protocol (v1.0, 26 September 2025) and will be finalized before access to full study data. Updates to the SAP will be versioned and aligned with protocol amendments. All SAP versions will be archived in OSF with a change log; any deviations in analyses will be explicitly reported in publications as “post-hoc” or “exploratory”.

##### Statistical Expertise

This SAP was developed jointly by the Institute of Medical Informatics (RWTH Aachen) and IPHAM (University of Magdeburg), combining methodological and domain expertise in acute and emergency medicine.

### Overview

Urgent and emergency care in Germany is delivered across multiple, loosely connected sectors. In the absence of coherent, time-resolved data on patient movements between Emergency Medical Services (EMS), out-of-hours primary care (OOH), emergency departments (EDs), and inpatient care, inefficiencies and coordination gaps remain difficult to quantify. A process-centred, trans-sectoral analysis is required to characterise real-world patient pathways and identify actionable levers for improvement. The study aims to reconstruct, model, and analyse patient pathways for urgent health complaints across all relevant sectors of the healthcare system in a German model region.

We will employ a mixed-methods observational study design. Routine data from EMS, out-of-hours primary care, EDs, and subsequent inpatient care will be pseudonymized at source, linked via a trusted third party, and analysed within a trusted research environment. Time-stamped event logs will support process mining for discovery, conformance, and performance analysis alongside descriptive statistics with stratification by context, such as setting, time of day, and urgency. Anonymous cross-sectional surveys of patients and frontline professionals, complemented by quarterly snapshot surveys in out-of-hours primary care and interviews, will provide convergent evidence on motives, barriers, and coordination of utilisation behaviour. Enrolment for surveys is anticipated from the 4th quarter of 2025 for one year; routine data capture covers 1 January–31 December 2026; analyses and dissemination run through 31 December 2027.

##### Research Objectives

The primary objectives are the five top-level protocol questions (RQ1–RQ5). All numbered sub-items (e.g., RQ1.1–RQ1.9, RQ2.1–RQ4.5) are secondary extensions that operationalise additional analyses using the linked routine data, surveys, and process-mining outputs.

##### Sample Size Justification

This study uses existing routine data and large-scale surveys. No formal sample size calculation was conducted as this is an observational mixed-methods design. Precision will be evaluated descriptively, and sensitivity analyses will assess robustness.

##### Definitions of Populations to be Analysed

- Routine Data Population: all pseudonymised patients included from EMS, ED, out-of-hours, and inpatient data sources.
- Survey Population: patients and providers completing the questionnaire.
- Stakeholder Interviews: experts contributing qualitative data for de jure process models.
- Analysis subsets: Defined by episode windows (72h, exploratively longer) and complaint clusters (CEDIS, ICD-10).

#### Endpoints

##### Primary Endpoint

Quantitative description of trans-sectoral patient pathways for adult patients (≥18 years) with acute complaints. Pathways are operationalised as time-ordered sequences of urgent/emergency encounters within 72 hours or shared complaint clusters.

##### Secondary Endpoints

- Alignment (concordance) between reconstructed pathways from routine data and self-reported pathways (patients/providers).
- Frequency and features of inefficient or avoidable utilisation (e.g., repeated emergency contacts; use of high-acuity services for low-urgency complaints; bypassing coordination structures such as 116117).
- Characterisation of non-traumatic pain-related pathways (prevalence, transitions, delays).
- Reported barriers to access and coordination from surveys, analysed in relation to pathway characteristics.
- Methodological: accuracy of record linkage and performance of the data management infrastructure (e.g., match rate, sensitivity, specificity, PPV).
- Comparative process/performance indicators across contexts (e.g., urban vs. non-urban; time of day; sector of initial contact), including inter-event intervals and bottlenecks.

- Conformance of data-driven process models to expert-elicited (“de jure”) models (fitness, precision, F1, generalisation).
- Identification and description of pathway variants associated with repeated use or chronic utilisation patterns (>30 days).

##### Exploratory Endpoints

- Sensitivity of pathway/episode definitions to alternative temporal windows beyond 72 h (e.g., 24 h, 7 days, 30 days).
- Characterisation of paediatric (<18 years) urgent/emergency pathways

#### Statistical Analyses

Analyses will follow exploratory health services research standards. Statistical significance will be set at $p < 0.05$ (two-sided). Effect sizes and 95% CIs will be reported alongside p-values.

##### Quantitative Descriptive Statistics

Descriptive statistics will be used to characterise the study population, individual encounters, and aggregated care episodes. All analyses will follow conventional standards for exploratory health services research. Where appropriate, stratifications will be applied by care sector, demographic subgroups, presenting complaint, urgency level, and care context (e.g., urban vs. non-urban). Demographic and clinical variables will include age, sex, presenting complaints, triage categories, and urgency level. Urban versus non-urban residence will be defined based on postal code and population density, using thresholds from the German Federal Statistical Office or comparable classifications.

Encounters will be described by type, volume, and temporal distribution. Continuous variables will be summarised according to their distribution: for approximately normally distributed variables, mean and standard deviation will be reported; for skewed distributions, median and interquartile range will be used. Categorical variables will be summarised as absolute counts and relative frequencies with percentages. Distributions will be visualised using histograms, cumulative distribution functions, and density plots. For patient pathways, Sankey diagrams will be used to illustrate sectoral transitions in predefined key pathways, such as episodes beginning with an EMS deployment or hotline contact, and pathways of patients with frequent or repeated encounters.

Between-site variation and clustering effects will be addressed by applying multilevel modelling where appropriate, with model structure adapted to the nesting of encounters within providers and regions. Design effects will be reported to contextualise the extent of clustering. Depending on the outcome type, hierarchical models will be specified accordingly. Linear models will be used for continuous outcomes, logistic models for binary outcomes, negative binomial or zero-inflated models for count data, and frailty models for time-to-event outcomes. In contexts with very few clusters, fixed effects are preferred, whereas random effects are used when many units are available (e.g., practices or EMS deployments). Robust standard errors will be reported as sensitivity analyses to ensure stability of inference under alternative assumptions.

#### Process-Modeling

The collected data will be analysed with a focus on care pathways and their underlying process structures using process mining methodology. Depending on scope and feasibility, various event logs will be created, including sector-specific logs as well as event logs from combined datasets to capture duplicates, multiple encounters, or inappropriate utilisation. In addition, data-driven definitions of duplicates, multiple encounters, and inappropriate utilisation will be derived to ensure consistent characterisation across sectors.

Each event log will undergo a process-oriented descriptive analysis and visualisation. Key descriptive process characteristics will include the number of events per patient pathway as a measure of pathway length, the number and type of sector transitions, inter-event time intervals (e.g., time from hotline contact to ED presentation), the distribution of activities across patient pathways, temporal patterns of activity occurrence (e.g., by hour of day and weekday), and the number and distribution of process variants defined as distinct sequences of activities. Dotted charts will be used to visualise temporal patterns across cases [1].

To complement these data-driven reconstructions, de jure process models will be created through qualitative interviews with domain experts from each sector. At least three experts per sector will be interviewed individually using a semi-structured format to elicit knowledge about pathway starts, intermediate steps, decision points, and endpoints. These preliminary sectoral models will then be discussed in joint sessions with experts across all sectors to examine linkages and integration points. Interviews and joint sessions will be recorded and then transcribed for evaluation.

On the event log, process discovery techniques will be applied, including Alpha Miner [2], Alpha+ [3], Alpha++, Inductive Miner[4], Inductive Miner Infrequent[5], Heuristic Miner[6], and Region-Based Mining[7-9]. The resulting models will be evaluated by conformance checking using established metrics: fitness (alignments or token-based replay)[10-12], precision (escaping edges)[12 13], F1-score as the harmonic mean of fitness and precision, generalisation (replay frequency)[12], and simplicity (graph size and complexity)[12]. Conformance checking will also be applied to the de jure models derived from expert interviews.

The best-performing models will be ranked according to F1-score, followed by generalisation and simplicity, and the top ten candidates will be presented to domain experts. In group interviews, experts will be asked to assess the plausibility of these models, including comparison with the de jure model where applicable, and to discuss

discrepancies between modelled and real-world processes. Because some models are expressed in Petri nets or Business Process Model and Notation (BPMN), they will be converted into human-readable diagrams for expert review. The discussion will focus on which model best reflects actual practice and whether modelled behaviours, such as parallel activities, are feasible within operational constraints. Based on this feedback, the top 10 most promising models selected by the experts will be selected for further analysis by a majority vote of the participating experts.
Performance analyses will then be conducted on the selected models, calculating alignments, action durations, and inter-event intervals to identify potential bottlenecks. Where process models reveal choices or branching behaviour, decision mining[1] will be applied to explore the factors underlying divergent patient pathways. Decision-tree learning methods will be used to ensure interpretable results, which will subsequently be validated in discussions with domain experts.
Finally, comparative analyses will examine whether processes differ systematically across groups or contexts. Differences in activity occurrence or execution times will be tested between age groups, urban versus rural settings, and other relevant characteristics. Inferential statistics will be applied for statistical comparisons[14].

#### Analysis of Survey Data

After removing incomplete surveys, descriptive statistics will be used to summarise key variables, including frequencies, means, medians, standard deviations, and interquartile ranges. Stratified analyses will be performed for selected subgroups (by age, gender, perceived urgency, or reason for attendance) to identify potential variations in utilisation patterns. Correlation analyses will be performed to examine associations between continuous or ordinal variables (e.g., age, perceived urgency, frequency of ED visits). Pearson's correlation will be used for normally distributed, linear data, while Spearman's rank correlation will be applied for ordinal variables or non-normally distributed data. To examine associations between sociodemographic characteristics, health-related behaviour, and patterns of emergency department use, bivariate analyses (*Chi-square tests*, *independent t-tests*) will be applied. Effect sizes will be reported alongside p-values, using Cramer's V or Phi for categorical associations, Cohen's d for mean differences, and Pearson's r or Spearman's ρ for correlations, each with 95% confidence intervals. Conventional benchmarks will be applied for interpretation while also considering contextual and clinical relevance. If emerging patterns suggest meaningful differentiation within the patient population, a cluster analysis (hierarchical or k-means clustering) will be performed to identify potential subgroups with distinct utilisation profiles. Furthermore, we will explore the feasibility of constructing a multivariable regression model to identify potential associations between patient characteristics and patterns of ED utilisation. In multivariable regression analyses, effect sizes will be expressed as odds ratios (OR) or regression coefficients with 95% confidence intervals.

Given that no linkage to routine clinical or hospital data is available, all analyses will be limited to self-reported variables. The suitability of this modelling approach will depend on the distribution, completeness, and variability of relevant survey items. If appropriate, regression analyses will be conducted to examine potential predictors of frequent ED attendance or non-urgent utilisation, using available sociodemographic and behavioural variables. All statistical tests will be two-sided, with a significance level set at $p < 0.05$.

Questionnaires for stakeholders will include open-ended questions, allowing them to provide input beyond the predefined answers suggested by the research team. Those responses will be analysed using qualitative content analysis, applying an inductive coding approach to identify recurring themes and structural issues.

#### Mixed-Methods Integration

To integrate quantitative routine data with survey-based insights, we will apply a joint display approach that enables the structured triangulation of observed patient pathways and self-reported experiences. Routine data will provide reconstructed, time-resolved pathways, while survey data will contribute subjective accounts of navigation, decision-making, and perceived barriers. Integration will be guided by predefined thematic categories (e.g., timing, access, redirection, repeat use) and stratified by patient characteristics and care contexts. Joint displays will align pathway patterns with reported motives and experiences, enabling cross-validation and the identification of mismatches between documented processes and perceived care. Integration will follow a convergence model, highlighting agreement, complementarity, or dissonance between data sources. Analyses will focus on cases and subgroups with available information from both sources, without requiring individual-level linkage. Results will inform the interpretation of structural findings and support the development of patient-centred steering strategies.

#### Subgroup Analyses

All primary and secondary analyses will be stratified by relevant demographic, clinical, and contextual variables, including age, sex, urgency, population density, time of presentation, Elixhauser comorbidity index[15], and sector of initial contact. Subgroup analyses will examine whether process characteristics, pathway variants, or survey-reported experiences differ systematically across these strata. A dedicated subgroup analysis will focus on patients presenting with pain as the leading symptom, given the clinical relevance, frequency, and potential for misdirection in this group. The cohort will be identified based on CEDIS chapter classifications, triage diagrams, and ICD-10 codes. For this subgroup, we will assess patient pathways, delays, sector transitions, and self-reported barriers in relation to documented process metrics. Interaction effects will be tested for key variables where effect modification is plausible, particularly for repeated use or bypassing coordination structures. Sensitivity analyses will be conducted to assess the impact of missing data (e.g., underreporting in specific sectors or incomplete timestamps) and to evaluate the robustness of the findings under varying assumptions regarding data completeness and dropout mechanisms.

In addition to these predefined strata, further subgroup analyses will be conducted for clinically or organizationally relevant conditions, care sectors, and provider specialities. These subgroups will be defined by presenting complaints, discharge diagnoses, or sector-specific utilisation patterns. To identify relevant disease and complaint clusters, we will apply a combined strategy: (i) mapping to standardised classification systems such as ICD-10 discharge diagnoses and CEDIS presenting complaints, (ii) grouping by triage categories (e.g., Manchester Triage System), and (iii) data-driven clustering of high-frequency or high-impact complaints using descriptive frequency analysis and clinical expert review. For each identified subgroup, dedicated process models will be constructed to visualise pathway structure, quantify throughput times, and characterise sector transitions.

#### Interim Analyses

Interim analyses will be limited to descriptive summaries for quality assurance and plausibility checking. Final analyses will be performed after database lock according to the Statistical Analysis Plan.

#### Statistical Analysis Changes from Protocol

Any changes to methods described in the protocol will be documented and justified in the SAP revision history.

#### Conventions

To ensure consistency, reproducibility, and interpretability of all analyses, the following conventions will apply throughout the study:

- All primary analyses will be two-sided with $\alpha = 0.05$, unless otherwise specified.
- Effect sizes will be reported alongside p-values and 95% confidence intervals.
    - For categorical data: Cramer’s V or Phi;
    - For mean differences: Cohen’s d; for correlations: Pearson’s r or Spearman’s ρ;
    - For regression: odds ratios or regression coefficients. Benchmarks: small ($d \approx 0.2$, $r \approx 0.1$, $V \approx 0.1$), moderate ($d \approx 0.5$, $r \approx 0.3$, $V \approx 0.3$), large ($d \geq 0.8$, $r \geq 0.5$, $V \geq 0.5$).
- All descriptive analyses will include counts (n) and denominators. Percentages will be calculated on non-missing data unless otherwise specified.
- Secondary and subgroup analyses will be interpreted with caution and with respect to multiple testing; effect sizes and confidence intervals will be reported.

- Where appropriate, the false discovery rate (FDR, Benjamini–Hochberg) will be explored as a sensitivity approach to assess the robustness of signals under multiplicity adjustment.
- Results from exploratory subgroup analyses will be explicitly labelled as such and not overinterpreted as confirmatory evidence.

#### Derived Variables

Encounter: A documented contact with any urgent or emergency care service (112 EMS hotline, 116117 hotline, EMS deployment, out-of-hours primary care, hospital ED, or GP/specialist coded as emergency). Minimum data required: patient pseudonym, date, and provider/sector.

Patient Pathway: A patient's ordered sequences of encounters within an episode, spanning sectors and providers:

Encounters re-occurring within 72 hours *or* the same main complaint cluster (CEDIS category or ICD-10 group) are grouped into the same episode; longer windows may be explored.

- A new episode is assumed if the interval *between two encounters* exceeds 72 hours *and* the main complaint cluster changes (CEDIS category or ICD-10 group).
- Episodes are capped at 30 days, in line with established urgent care indicators; longer sequences are considered chronic utilisation patterns.

Patient Pathways are characterised by:

- Number of encounters (events grouped into episodes).
- Sequence of sectoral transitions (activity sequence).
- Timing between events.
- Types of activities recorded in the event log.

Emergency Patient Pathway: Defined as the contiguous core segment of encounters addressing the urgent complaint, where gaps between encounters do not exceed 360 minutes.

#### Imputation of Dates

Missing data are an inherent characteristic of routine documentation and are expected to occur across all sectors and data sources. In contrast to controlled study environments, documentation in emergency and urgent care settings is not designed primarily for research purposes, and missingness is often structurally induced—for example, due to undocumented handovers, unavailable timestamps, or system-level constraints. As such, we do not assume data to be missing at random in the strict statistical sense. Instead, we acknowledge that missingness may itself reflect real-world process characteristics and operational limitations, and thus carries informative value. Therefore, rather than applying imputation or exclusion strategies that may introduce bias, missing values will be explicitly preserved in the data and analysed descriptively as part of the process characterisation. Sensitivity analyses will evaluate their impact. For time-critical process steps (e.g., timestamps, triage assessments), missingness will be reported per sector and subgroup, and sensitivity analyses will be conducted to evaluate the potential impact on pathway reconstruction and indicator derivation.

#### Software and Reproducibility

All analyses will be conducted using established statistical and process mining software (e.g. R, Python, PM4Py). Exact versions, packages, and libraries will be specified in publications. Code management will follow version control standards (e.g. GitHub), ensuring full reproducibility.

#### Reporting & Dissemination

Results will be reported according to established guidelines for observational and mixed-methods studies (STROBE, RECORD, COREQ for qualitative data). Findings will be disseminated through peer-reviewed publications, conference presentations, and structured summary reports for stakeholders.

### Appendix

#### Research Questions

*Table 1: Tentative Research Questions*

| RQ | Research question | Key operationalisation | Data & cohort | Primary methods |
|---|---|---|---|---|
| RQ1 (Primary) | How do adults with acute complaints move through urgent & emergency care—what are the main sequences and transitions? | Pathway variants, transition matrices, inter-event times, episode length, entry points (EMS, OOH, EDs, telemedicine); 72h linkage window; 30d cap; ≤360min emergency core segment | EHR Data, Survey Data, Snapshot Survey Data, Stakeholder Surveys, Interviews | Process mining, descriptives |
| RQ1.1 | Which pathway process models are discoverable and how representative are they? | Model fitness, precision, F1, variant entropy, stability by sector/time | EHR Data | Process mining |

| | | | | |
|---|---|---|---|---|
| RQ1.2 | How does EMS deployment relate to subsequent treatment up to ED handover? | Time to handover, destination (ED vs OOH), transitions, admission rate by vehicle/priority | EHR Data | Process mining, multivariable regression |
| RQ1.3 | How do normative (de jure) vs. data-driven pathway models compare? | Fitness, precision, plausibility vs expert models | EHR Data | Process mining |
| RQ1.4 | How do OOH-managed vs. ED-presenting pathways differ? | Variant prevalence, episode length, inter-event times, disposition, redirections, | EHR Data | Process mining, descriptives |
| RQ1.5 | Which pathway variants occur in both OOH and EDs? | Matched variants (activities/timing), outcomes (admission, revisit 72h/30d) | EHR Data | Process mining, survival analysis |
| RQ1.6 | Are there urban vs. non-urban differences in EMS use and destination? | EMS call rates, destinations, time metrics, adjusted differences, classified by postal code / population density | EHR Data | Process mining, multilevel models (patients nested in regions) |
| RQ1.7 | How do utilisation and pathway structures vary by time of day and OOH? | Variant prevalence, delays by hour/weekday, off-hours vs in-hours | EHR Data | Process mining, descriptives |
| RQ1.8 | How do pathways differ by age, sex, and demographics? | Variant mix, transition probabilities, outcomes by strata | EHR Data | Process mining, multivariable regression |
| RQ1.9 | Do sex-specific pathways deviate from normative models? | Sex-specific fitness/precision vs de jure models | EHR Data | Process mining |
| RQ1.10 | What role do tele-emergency physician system encounters play in acute care pathways, and how do these patients' pathways differ from those without such encounters? | Pathway variants, episode length, entry points (telemedicine vs EMS/OOH/ED) | EHR Data, Stakeholder Surveys | Process mining, descriptives |
| RQ1.11 | How do pathways of pediatric patients differ? | Pathway variants, episode length, entry points | EHR Data, Stakeholder Surveys | Process mining, descriptives |
| RQ2 (Primary) | To what extent does privacy-preserving linkage capture full trans-sector pathways, and what does this reveal for empowerment and efficiency? | Linkage completeness, pathway coverage, missing-step profiles; empowerment/efficiency indicators | EHR Data, Survey Data, Snapshot Survey Data, Stakeholder Surveys, Interviews | Process mining, linkage metrics |
| RQ2.1 | What share of EMS-initiated low-acuity cases receive ED care? | Proportion ED care, admission rate, revisit 72h, MTS/ESI Blue Green | EHR Data Survey Data, Snapshot Survey Data | Multivariable regression, descriptives |
| RQ2.2 | Among ED patients, which could have been ambulatory-managed or telemedicine-managed? | Low-acuity, discharged, no inpatient ≤7–30d; misalignment rate | EHR Data Survey Data, Snapshot Survey Data | Multivariable regression, rules-based classification |
| RQ2.3 | How can process mining models inform resource management in | Candidate levers (staffing, routing thresholds, telemedicine options); simulated impact | EHR Data, Survey Data, Interviews | Process mining, |

| | | | | |
|---|---|---|---|---|
| | acute care and guide actionable recommendations? | | | Qualitative Evaluation |
| RQ2.4 | Are there sex differences in low-acuity contacts and treatment? | Rate ratios, diagnostics/interventions, revisit, treatment intensity, MTS/ESI Blue Green | EHR Data Survey Data, Snapshot Survey Data | Multivariable regression, descriptives |
| RQ2.5 | Do triage results and subsequent decisions differ by sex? | Distribution of triage levels, disposition, resource use | EHR Data Survey Data, Snapshot Survey Data | Multivariable regression, descriptives |
| RQ2.6 | What proportion adheres to 116117/112/telemedicine recommendations? | Adherence (recommended vs realised care), predictors (age, sex, timing, complaint) | EHR Data Survey Data, Snapshot Survey Data | Multivariable regression |
| RQ3 (Primary) | Which patient/provider perspectives and barriers are linked to inefficient or avoidable use, and how can they inform empowerment? | Health literacy, motives, barriers; linked to pathway indicators (redirections, revisits, avoidable ED proxy, telemedicine uptake) | Survey Data, Snapshot Survey Data, Stakeholder Surveys, Interviews | Multivariable regression, mixed-methods integration |
| RQ3.1 | Which perspectives most strongly predict inefficient use? | Associations of survey scales with pathway indicators | Survey Data, Stakeholder Surveys | Multivariable regression, Qualitative Evaluation |
| RQ3.2 | Which pathway patterns indicate structural or informational barriers? | Redirections, waits, dead-ends; barrier taxonomy | Survey Data, Snapshot Survey Data | Process mining, Qualitative Evaluation |
| RQ3.3 | Can process models support adaptive recommendations? | Feasibility of real-time rules, projected effects (including telemedicine options) | EHR Data,<br><br>Survey Data, Stakeholder Surveys | Process mining, |
| RQ3.4 | What sex-specific barriers affect navigation and help-seeking? | Misrouting/non-adherence by sex | Survey Data, Stakeholder Surveys | Multivariable regression |
| RQ3.5 | How do genders perceive navigation roles and empowerment strategies? | Preferred info channels, perceived usefulness, uptake intent (including telemedicine) | Survey Data, Stakeholder Surveys | Descriptives, Qualitative Evaluation |
| RQ4 (Primary) | What patterns indicate duplicate, multiple, or inappropriate utilisation, and which variants carry higher risk? | Repeat use (72h/30d), redundant activities, misdirection markers, utilisation intensity | EHR Data, Survey Data, Snapshot Survey Data, Interviews | Process mining, multivariable regression |
| RQ4.1 | Which variants signal multiple or redundant actions? | Duplicated diagnostics/referrals | EHR Data, Interviews | Process mining, Qualitative Evaluation |
| RQ4.2 | Which deviations or repetitions identify duplicates/errors? | Repetition motifs, deviation from reference models | EHR Data, Interviews | Process mining, Qualitative Evaluation |

| | | | | |
|---|---|---|---|---|
| RQ4.3 | Do specific sequences or delays trigger double requests? | Triggers such as long dispatch-to-handover | EHR Data, Interviews | Process mining, time-to-event, Qualitative Evaluation |
| RQ4.4 | Are there sex-specific patterns in multiple utilisation? | Sex-stratified repeat use rates | EHR Data, Interviews | Multivariable regression, descriptives, Qualitative Evaluation |
| RQ4.5 | Is it possible to predict 72h revisits with similar complaints? | Predictive accuracy for revisits | EHR Data | Process mining, Machine learning, Multivariable regression |
| RQ5 (Primary) | What is the significance of non-traumatic pain as a pathway phenotype, and where are improvement opportunities? | Non-traumatic pain (CEDIS/ICD-10, >24h); share of daily load; routing appropriateness | EHR Data, Survey Data,, Interviews | Process mining, descriptives |
| RQ5.1 | How prevalent are non-traumatic pain pathways? | % of urgent volume, temporal patterns | EHR Data | Descriptives |
| RQ5.2 | To what extent do non-traumatic pain episodes route to ambulatory/telemedicine, out-of-hours care, ED, or EMS, and which factors are associated with ED routing? | ED low-intensity with no admission nor diagnostics short LOS; Ambulatory or telemedicine but no inpatient ≤7d; EMS low-acuity ED without admission; escalation from ambulatory to ED/hospital ≤72h; safety ≤7d return admission. | EHR Data, Interviews, Survey Data | Multivariable regression, rules-based classification, Qualitative Evaluation |
| RQ5.3 | How often do pain cases escalate to inpatient ≤7d and what predicts escalation? | 7d admission, predictive signals (timing, entry, triage, telemedicine, involvement) | EHR Data | Multivariable regression, survival analysis |
| RQ5.4 | Which empowerment opportunities emerge from pain-pathway models? | Candidate messages/rules, projected impact | Survey Data, Stakeholder Surveys | Scenario testing, Qualitative Evaluation |

#### Data Dictionary

##### *116117 medical hotline (telephone triage)*

| Attribute | Type |
|---|---|
| call_ts | DATETIME |
| sex | STRING |
| street | STRING |
| zip_code | STRING |
| tts_med | STRING |
| poc_smed | STRING |
| call_reason | STRING |
| secondary_complaints | STRING |
| poc_disposition | STRING |
| disposition_ts | DATETIME |

##### *Outpatient & out-of-hours primary care*

| Attribute | Type |
|---|---|
| datetime | DATETIME |
| sex | STRING |
| age | INT |
| zip_code | STRING |
| specialty_group | STRING |
| event_type | STRING |
| services | STRING |
| diagnoses_icd | LIST<STRING> |

##### *Provider facility reference*

| Attribute | Type |
|---|---|
| hospital_id | STRING |
| practice_id_bsn | STRING |
| physician_id_lanr | STRING |
| specialty_group_code_fgc | STRING |
| practice_postal_code | STRING |

##### *Emergency Medical Services (EMS)*

| Attribute | Type |
|---|---|
| age | int |
| mission_type | STRING |
| mission_keyword | STRING |

| | |
|---|---|
| mission_keyword_text | STRING |
| call_ring_ts | DATETIME |
| call_answer_ts | DATETIME |
| incident_country | STRING |
| incident_state | STRING |
| incident_region | STRING |
| incident_district | STRING |
| incident_city | STRING |
| incident_street | STRING |
| incident_object | STRING |
| destination_country | STRING |
| destination_state | STRING |
| destination_region | STRING |
| destination_district | STRING |
| destination_city | STRING |
| destination_city_part | STRING |
| destination_street | STRING |
| destination_object | STRING |
| diagnosis_text | STRING |
| unit_name | STRING |
| unit_type | STRING |
| dispatch_ts | DATETIME |
| bos_status1_available_radio_ts | DATETIME |
| bos_status2_available_station_ts | DATETIME |
| bos_status3_enroute_ts | DATETIME |
| bos_status4_on_scene_ts | DATETIME |
| bos_status7_patient_onboard_ts | DATETIME |
| bos_status8_at_destination_ts | DATETIME |

*Emergency Department (ED)*

| Attribute | Type |
|---|---|
| hospital_id | STRING |
| arrival_ts | DATETIME |
| triage_ts | DATETIME |
| triage_system | STRING |
| triage_acuity | STRING |
| cedis_presenting_complaint | STRING |
| physician_contact_ts | DATETIME |
| lab_ts | DATETIME |
| blood_gas_ts | DATETIME |
| ekg_ts | DATETIME |
| urine_test_ts | DATETIME |
| ultrasound_ts | DATETIME |
| echocardiography_ts | DATETIME |
| emergency_ct_head_ts | DATETIME |
| ct_ts | DATETIME |
| trauma_scan_ts | DATETIME |
| xray_spine_ts | DATETIME |
| xray_chest_ts | DATETIME |
| xray_pelvis_ts | DATETIME |
| xray_extremities_ts | DATETIME |
| mri_ts | DATETIME |
| therapy_start_ts | DATETIME |
| discharge_ts | DATETIME |
| mode_of_arrival | STRING |
| referral_source | STRING |
| isolation_status | BOOLEAN |
| isolation_reason | STRING |
| disposition (e.g., home, ward, ICU, transfer, death) | STRING |
| sex | STRING |
| age_years | INTEGER |
| patient_postal_code | STRING |
| ed_diagnoses_icd10 | LIST<STRING> |
| ed_leading_diagnose_icd10 | STRING |
| ed_additional_codes_icd10 | LIST<STRING> |

##### *Inpatient care — admissions, procedures, transfers*

| Attribute | Type |
| --- | --- |
| hospital_id | STRING |
| inpatient_admission_ts | DATETIME |
| admission_reason_code | STRING |
| admission_type_code (E,Z,N,R,V,A,G,B) | STRING |
| case_merge_flag | BOOLEAN |
| case_merge_reason_code | STRING |
| icu_los_days | INTEGER |
| ventilation_hours | INTEGER |
| postinpatient_end_date | DATE |
| postinpatient_days | INTEGER |
| specialty_transfer_ts | DATETIME |
| specialty_admission_ts | DATETIME |
| specialty_discharge_ts | DATETIME |
| icu_bed_flag | BOOLEAN |
| procedure_ops_ts | DATETIME |
| ops_version | STRING |
| ops_code | STRING |
| inpatient_discharge_ts | DATETIME |
| discharge_reason_code | STRING |
| diagnosis_type (HD/ND) | STRING |
| icd_version | STRING |
| icd_code | LIST <STRING> |
| diagnostic_certainty (A/V/Z/G) | STRING |
| sex | STRING |
| age_years | INTEGER |
| patient_postal_code | STRING |

*Hashed Identifiers stored separately, used only for privacy-preserving record linkage*

| Attribute | Type |
|---|---|
| first_name | STRING |
| last_name | STRING |
| date_of_birth | DATE |
| health_insurance_number | STRING |
| health_card_number_egk | STRING |
| phone_number | STRING |
| street_address | STRING |
| patient_postal_code_full | STRING |
| hospital_patient_number (as stored; or hashed with salt) | STRING |
| hospital_case_number (as stored; or hashed with salt) | STRING |
| ems_incident_number | STRING |
| control_centre_call_id | STRING |
| national_insurer_ik_number | STRING |
